# Decline in prevalence of tuberculosis following an intensive case-finding campaign and the COVID-19 pandemic in an urban Ugandan community

**DOI:** 10.1101/2023.03.03.23286745

**Authors:** Emily A Kendall, Peter J Kitonsa, Annet Nalutaaya, Katherine O Robsky, Kamoga Caleb Erisa, James Mukiibi, Adithya Cattamanchi, Midori Kato-Maeda, Achilles Katamba, David W Dowdy

## Abstract

**Background:** Systematic screening is a potential tool for reducing the prevalence of tuberculosis and counteracting COVID-related disruptions in care. Repeated community-wide screening can also measure changes in the prevalence of tuberculosis over time.

**Methods:** We conducted serial, cross-sectional tuberculosis case-finding campaigns in one community in Kampala, Uganda, in 2019 and 2021. Both campaigns sought sputum for tuberculosis testing (Xpert MTB/RIF Ultra) from all adolescents and adults. We estimated the prevalence of tuberculosis among screening participants in each campaign and compared characteristics of people with tuberculosis across campaigns. We simultaneously enrolled and characterized community residents who were diagnosed with tuberculosis through routine care and assessed trends in facility-based diagnosis.

**Results:** We successfully screened 12,033 community residents (35% of the estimated adult/adolescent population) in 2019 and 11,595 (33%) in 2021. In 2019, 0.94% (95% CI 0.77-1.13%) of participants tested Xpert-positive (including trace). This proportion fell to 0.52% (95%CI 0.40-0.67%) in 2021; the prevalence ratio was 0.55 [95%CI: 0.40-0.75]). There was no change in the age (median 26 vs 26), sex (56% vs 59% female), or prevalence of chronic cough (49% vs 54%) among those testing positive. By contrast, the rate of routine facility-based diagnosis remained steady in the eight months before each campaign (210 [95%CI 155-279] vs. 240 [95%CI 181-312] per 100,000 per year).

**Conclusions:** Following an intensive initial case-finding campaign in an urban Ugandan community in 2019, the burden of prevalent tuberculosis as measured by systematic screening had decreased by 45% in 2021, despite the intervening COVID-19 pandemic.

## Introduction

The COVID-19 pandemic disrupted systems of diagnosis and treatment for tuberculosis (TB). TB notifications worldwide fell by 18% in 2020 relative to 2019, and recovered only partially in 2021 [1]. The prevailing explanation for this decline is a reduction in case finding and diagnosis [2–5], and models estimate that reductions in transmission are unlikely to have completely mitigated this negative impact [6,7]. As such, global TB incidence, prevalence, and mortality are projected to have increased in 2020 and 2021 for the first time in over two decades [1,7–9].

There is potential, however, to restore or even accelerate pre-pandemic declines in TB incidence, prevalence, and mortality, by undertaking interventions to find and treat more people with TB. Intensive case-finding efforts were shown to reduce the community prevalence of TB in certain pre-COVID clinical trials [10,11], and models estimate that case-finding can also overcome adverse effects of the COVID pandemic on TB burden [6,12]. Thus, it is possible that, in the context of ambitious TB case finding, TB prevalence could fall substantially from pre-pandemic levels.

Formal prevalence surveys have not yet directly compared the prevalence of TB before versus after the pandemic (with the exception of India’s national survey, which did not include pre/post measurements in the same regions [13]). Community-based active case-finding efforts may also be used to measure relative changes in the burden of prevalent TB, if conducted serially with consistent methodology. However, intensive efforts to find prevalent TB may also reduce the burden they are measuring, as the people who are found with TB are linked to treatment and no longer contribute to prevalence or transmission [10]. Thus, changes in the yield of serial active case-finding campaigns reflect the combined effects of external events (e.g. other health interventions and secular trends) and case-finding itself.

In 2019, we undertook a community-wide campaign of active TB case finding in Kampala, in the context of intensive community-based case finding and any potential impacts of the COVID-19 pandemic response Uganda, as part of an effort to characterise all individuals TB within a single community [14]. In 2021, we conducted a second campaign using the same methods. We report how the prevalence and characteristics of TB among participants changed in the aftermath of our initial campaign and the intervening COVID pandemic.

## Methods

### Study design and population

We performed two population-wide TB screening campaigns in a single, spatially defined urban community (estimated adult/adolescent population 35,000; land area 2.2 km^2^) in Kampala, Uganda. The first campaign occurred from February 4, 2019, through November 30, 2019 (300 days), and the second occurred from February 3, 2021, through August 19, 2021 (198 days). Both campaigns used the same door-to-door and venue-based screening methods with the goal of screening as many adults and adolescents (age 15+ years) as possible throughout the entire community. All consenting adult/adolescent residents, irrespective of symptoms, were offered Xpert MTB/RIF Ultra testing (Cepheid, Inc., Sunnyvale, CA, USA) using expectorated sputum.

Additional study activities, which were not used to estimate TB prevalence among screening participants but which had potential to affect TB burden in the study community, included: enumeration of all community residents diagnosed with TB through routine care; recruitment of all patients routinely diagnosed with TB and all screening participants with M. tuberculosis detected by Xpert Ultra, for detailed investigations (see below); recruitment of selected Xpert-negative community residents for the same detailed investigations which included sputum culture; and contact investigation of the household and close non-household contacts of TB-positive individuals who enrolled into our study during either of the two campaigns. These activities are described in more detail below and in supplemental text 1.

### Community-based TB screening procedures

During each campaign, we systematically moved “door-to-door” through the community, visiting every residential and commercial building and offering sputum screening to every adult/adolescent encountered. During each campaign, we also held approximately ten venue-based screening events, generally on weekends after advertisement in neighboring zones.

Consenting participants with or without TB symptoms were coached and asked to provide the best expectorated sputum sample that they could; success in expectoration is described in Supplemental Text 5. Participants with positive Xpert Ultra results were linked to care by study staff, except for those with “trace” results during the second campaign, who were counted in our primary prevalence ratio estimate but evaluated further before recommending treatment (Supplemental Text 2).

As study staff moved through the community, they also enumerated each residence and asked household members or neighbors to report the number of adult/adolescent and child residents; these data were used for population estimates (Supplemental Text 3). To identify episodes of repeat contact with the study (e.g., during both campaigns), all screening participants underwent biometric iris scanning (iRespond, irespond.org, USA).

### Patients diagnosed at health facilities

We also enumerated and recruited all adult/adolescent community residents who were evaluated and diagnosed with TB in the community’s four outpatient TB diagnosis and treatment centers (a large public health center, a private HIV clinic, and two smaller medical clinics) from May 2018 through August 2021, using regular review of clinic registers and conversations with clinic staff.

### Detailed investigations of cases and controls

All adult/adolescent residents with a positive (including trace) Xpert screening result, as well as all residents diagnosed with TB through other routine or study-related activities, were recruited as “cases” for more detailed investigations. These additional procedures included a detailed questionnaire about TB history, symptoms, risk factors, and care seeking; additional expectorated sputum collection for liquid and solid mycobacterial culture;[15] blood C-reactive protein (CRP) measurement and HIV serological testing; contact investigation; and biometric iris scanning.

During the two campaigns, we also recruited one Xpert-negative “control” individual for each enrolled case. Controls were selected at random from the index individual’s zone of residence using a household-based sampling frame and asked to complete the same additional procedures as cases.

### Statistical methods

Our primary outcome was the crude TB prevalence ratio among community-based screening participants, comparing the second campaign to the first. We calculated the prevalence in each campaign as the proportion of valid Xpert screening results that were positive or trace. Secondary prevalence estimation is detailed in Table S1: (a) age- and sex-adjusted, (b) excluding Xpert-trace-positive/culture-negative cases, and (c) the proportion of the total study area population diagnosed with TB (including through contact investigation or routine care) during each campaign. Prevalence estimates are reported with 95% exact binomial confidence intervals (95%CIs). Adjusted and unadjusted prevalence ratios are estimated using log-binomial regression (R package logbin).

To estimate overlap between the populations participating in the two campaigns, we compared the proportion of iris IDs that appeared in both datasets against the proportion expected by random chance if the underlying population were unchanged, assuming also that missing identification numbers (for example due to technical malfunction, equipment unavailability, or participant refusal) were missing at random (details in Supplemental Text 6). These biometric estimates were used to estimate population turnover and compared to self-reported participation and migration histories.

We calculated rates of TB diagnosis in the study area, through facility-based and community-based case finding, during time intervals defined by the beginning and end of the study’s two active case-finding campaigns and by the beginning and end of a strict COVID-related lockdown (March – May 2020), during which nonessential public and private transport and most types of public and private gatherings were restricted and a 43% drop in regional notifications was reported [16]. The population denominator for each rate is the most proximal estimate of the community’s adult/adolescent population, and 95% confidence intervals are based on Poisson distributions.

Individual participant characteristics are summarized as proportions or as medians with interquartile ranges, and groupwise comparisons use chi-square tests with continuity correction or Kruskal-Wallis rank sum tests; missing results are indicated and excluded.

Study data were collected and managed using REDCap electronic data capture tools hosted at Johns Hopkins University [17,18]. All data analysis was performed using R version 4.1.0 (R Foundation for Statistical Computing, Vienna, Austria) [19].

### Ethical considerations

Participants gave verbal consent for TB screening and written informed consent for all additional procedures (assent and parental consent if <18 years of age). The study was approved by the Institutional Review Boards of the Johns Hopkins Bloomberg School of Public Health and Makerere University School of Public Health.

## Results

### Screened Population

A total of 12,033 study area residents (35% of the estimated adult/adolescent population) provided samples for screening and had valid Xpert results in 2019, and 11,595 (33%) were screened and had valid results in 2021 (Figure 1, Figures S1-S3). In addition, over the 39-month evaluation period from May 2018 to August 2021, an estimated 1,817 study area residents (5% of the estimated adult/adolescent population) were evaluated for presumptive TB at local health facilities.

**Figure 1:**
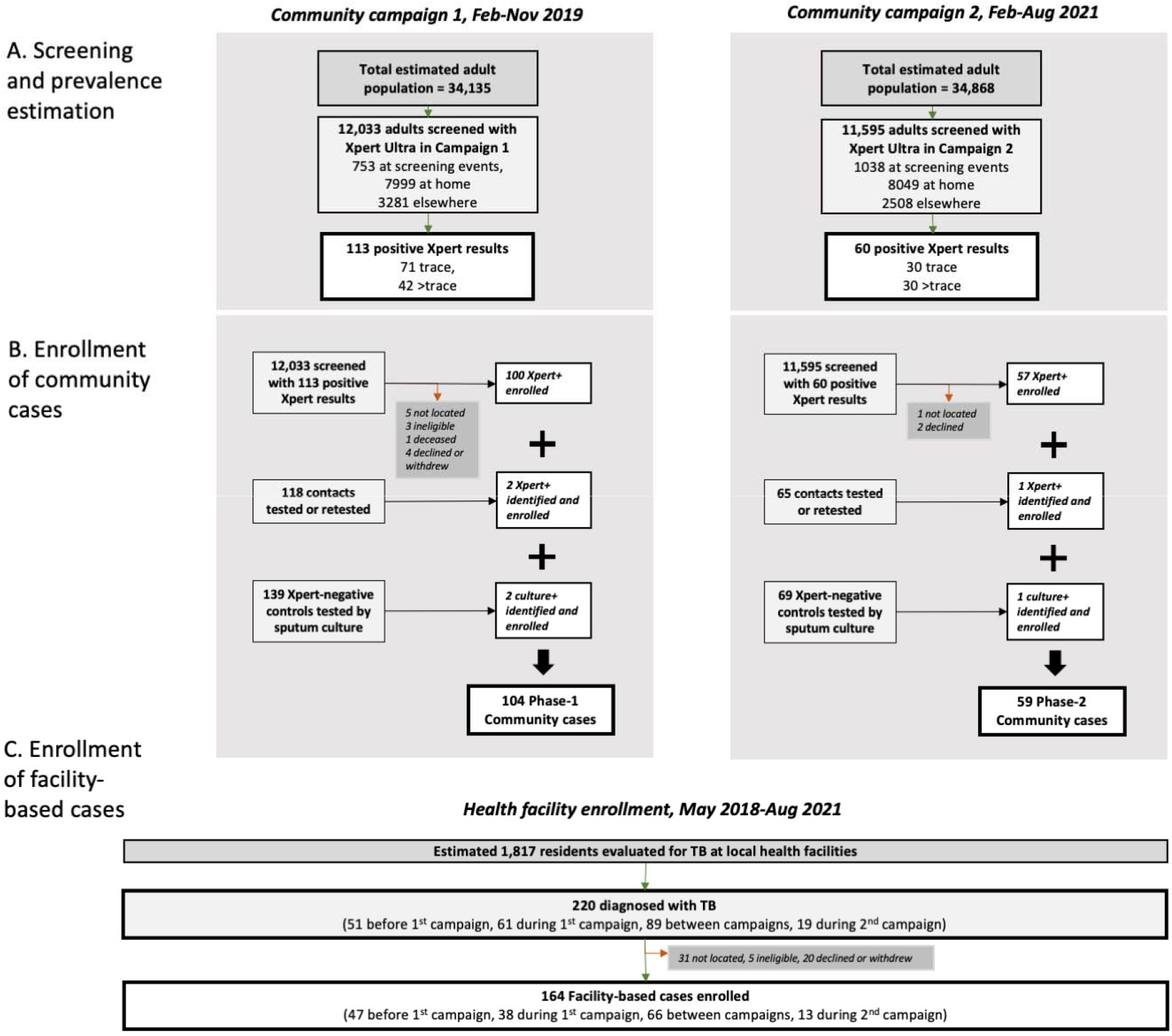
Flowchart of TB screening, community case enrollment, and enrollment of individuals diagnosed with TB at local health facilities. Prevalence estimates are based on screening in the community (Panel A). Additional investigations were then performed among those diagnosed with prevalent TB in the community, their household and non-household contacts, and matched individuals with negative Xpert results (“controls”) living in the same geographic zone (Panel B). From nine months before the first case-finding campaign to the end of the second campaign, individuals diagnosed with TB through routine care at four local health facilities were also enrolled (Panel C).

The age and sex compositions of the enumerated and participating populations were stable over time (Table 1, Figure S4). Participation by geographic zone and type of screening location were also consistent across the two campaigns (Table 1, Figure S5).

**Table 1:**
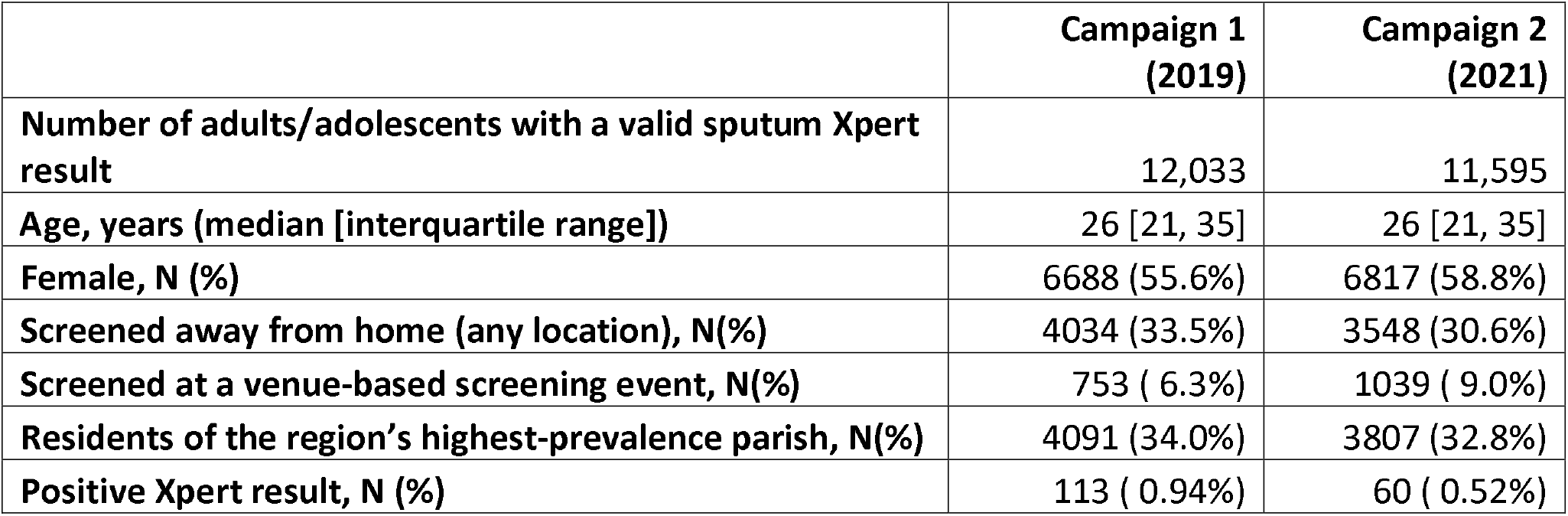
Characteristics of participants in two case-finding campaigns for tuberculosis in an urban Ugandan community.

Based on iris ID data, we estimated that 33-37% of the participants eligible for each campaign had participated in the other campaign as well (Supplemental Text 6 and Table S2).

### Estimates of TB Prevalence in the Screened Population

As reported previously [14], the prevalence of sputum Xpert-positive TB among participants in the first campaign was 0.94% (95% CI 0.77-1.13%; 113 of 12,033 valid results). In the second campaign, this prevalence fell to 0.52% (95% CI 0.40-0.67%; 60 of 11,595 valid results) (Figure 1). The TB prevalence ratio, comparing the second to the first campaign, was 0.55 (95% CI 0.40 - 0.75). TB prevalence also decreased under secondary definitions (Figure 2). TB prevalence among men fell from 1.22% to 0.61%, with a smaller decline among women from 0.72% to 0.45% (Figure S6), but the prevalence ratio remained stable (0.56, 95% CI: 0.41-0.76) after adjustment for age and sex. The prevalence ratio was also robust to adjustment by parish of residence and screening location (home versus other) (Table S3).

**Figure 2:**
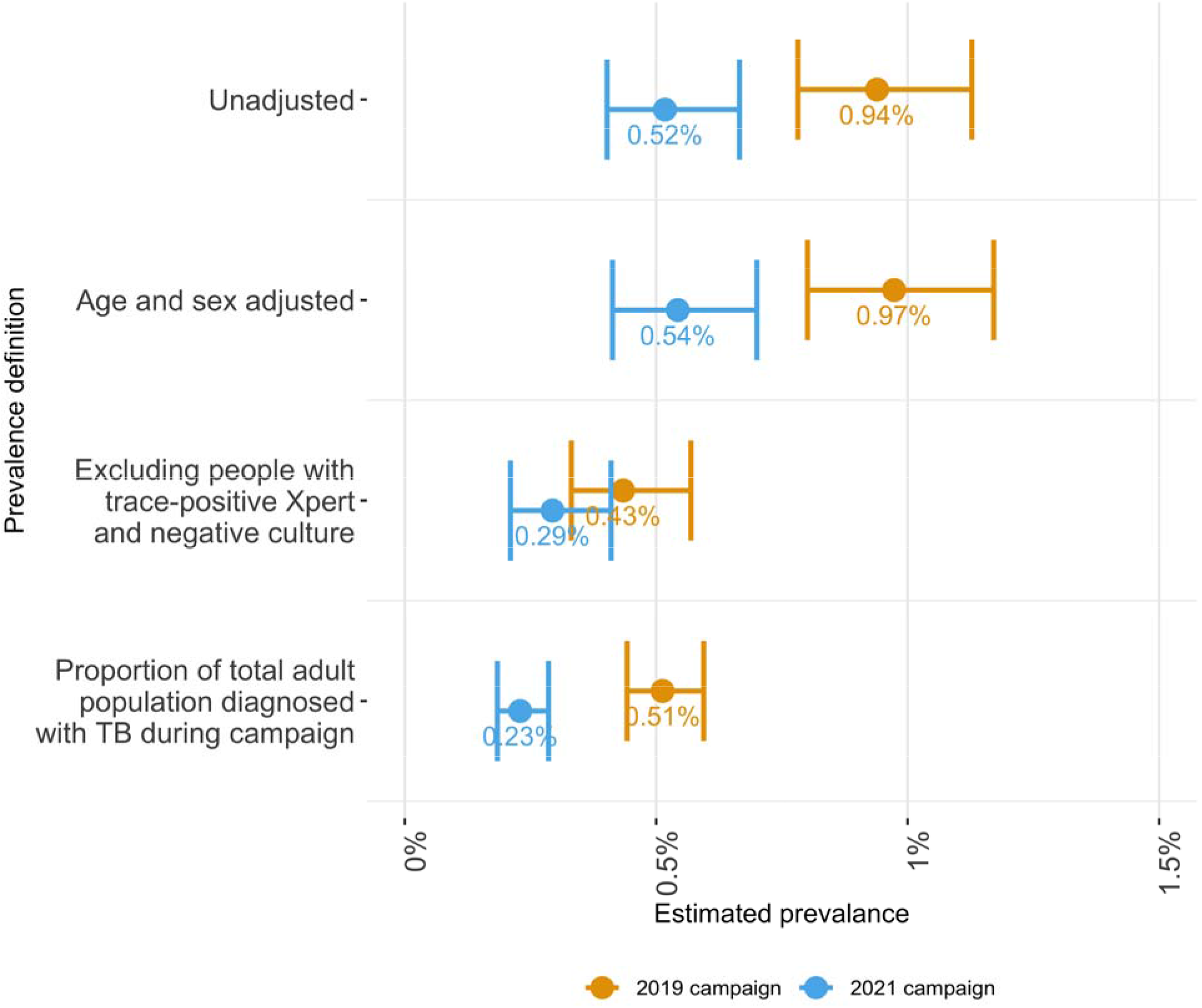
Prevalence of TB among community-based screening participants in 2019 and 2021, under primary and secondary definitions. Prevalence ratios with 95% confidence intervals are shown in italics. The first three estimates take as the denominator the number of people screened in the community and exclude individuals diagnosed through routine clinical care; the fourth estimate includes those diagnosed through routine care but uses the full estimated adult/adolescent population of the community as the denominator. Not included due to small sample size are estimates that include the prevalence of culture-positive TB among the small number of Xpert-negative individuals who underwent sputum culture (3 out of 208 individuals sampled across the two campaigns).

In the first campaign, 71 of 113 positive results (63%) were trace-positive, versus 30 of 60 (50%) in the second campaign. There was otherwise no consistent trend in the semiquantitative level of Xpert positivity between the two campaigns (Figure S7). When excluding trace-positive, culture-negative results from analysis, the TB prevalence ratio (comparing the second campaign to the first) was 0.68 (95% CI: 0.44 - 1.04).

Of 208 Xpert-negative community controls enrolled for more detailed investigations, 3 (1.4%) were diagnosed with TB by sputum culture (Figure 1).

### Concurrent Trends in Health Facility Notifications

The first community-based campaign accounted for 54% (87/161) of pulmonary TB diagnoses among adult/adolescent study area residents in 2019 (Figure 3).

**Figure 3:**
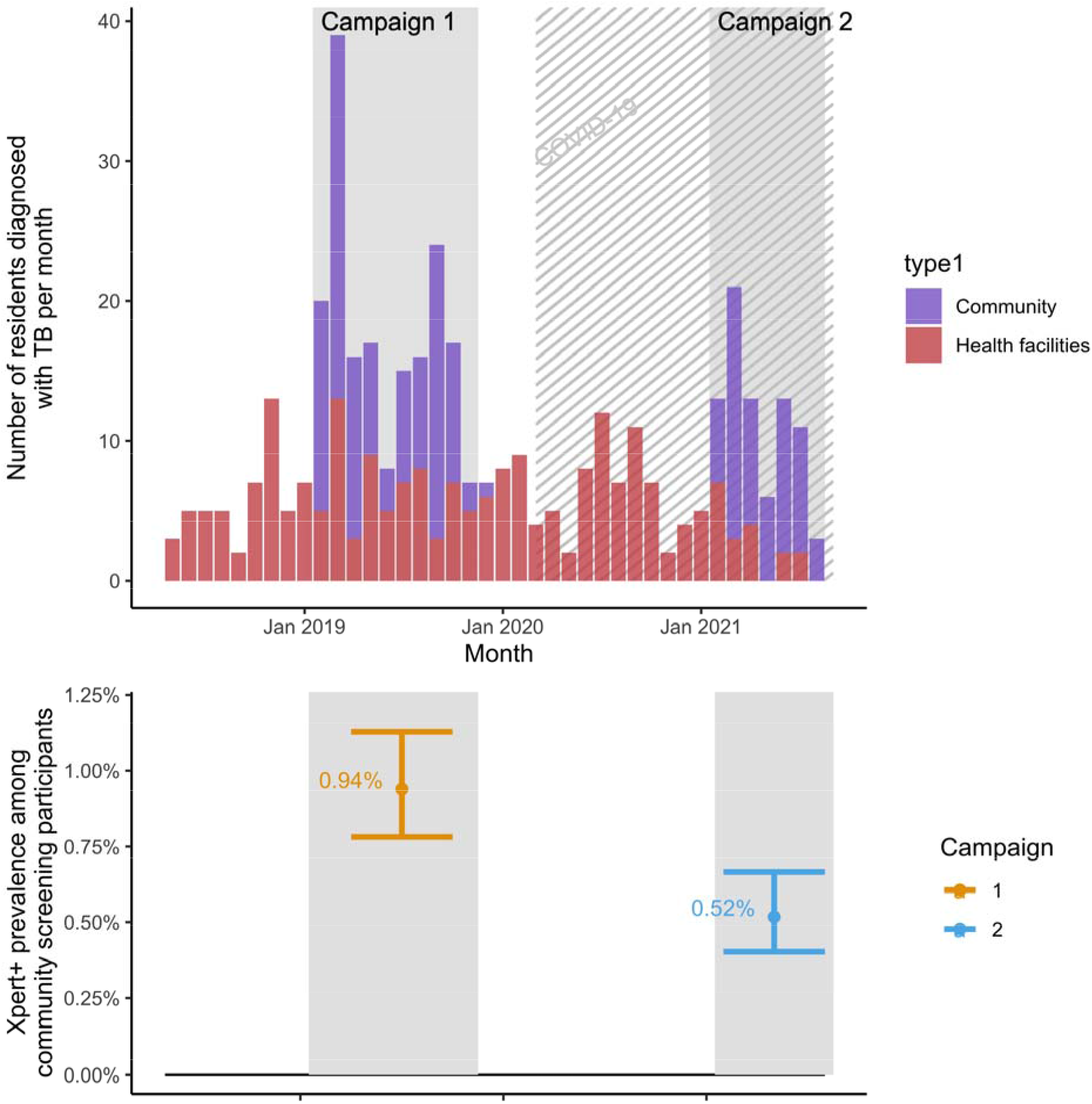
Timeline of community-based and facility-based TB diagnosis in an urban Ugandan community. Residents diagnosed through active case-finding activities (including community-wide screening and contact investigation) during the two case-finding campaigns are shown in purple as stacked bars, with routine TB notifications made in ambulatory care settings throughout the study shown in red. The COVID-19 pandemic is indicated with diagonal gray lines; local COVID-related restrictions and timelines are detailed in Supplemental text 4. The lower panel shows the timing of the primary prevalence estimates shown in Figure 2.

The rate at which study area residents were diagnosed with TB at health facilities decreased transiently at the start of the COVID pandemic, but was otherwise consistent over time from 2018 through 2020 (Table 3). The rate of routine TB diagnosis fell during the second active case-finding campaign, which also corresponded to a second COVID lockdown (Supplemental text 4) and use of the public health center’s TB clinical space for COVID vaccinations.

**Table 2:**
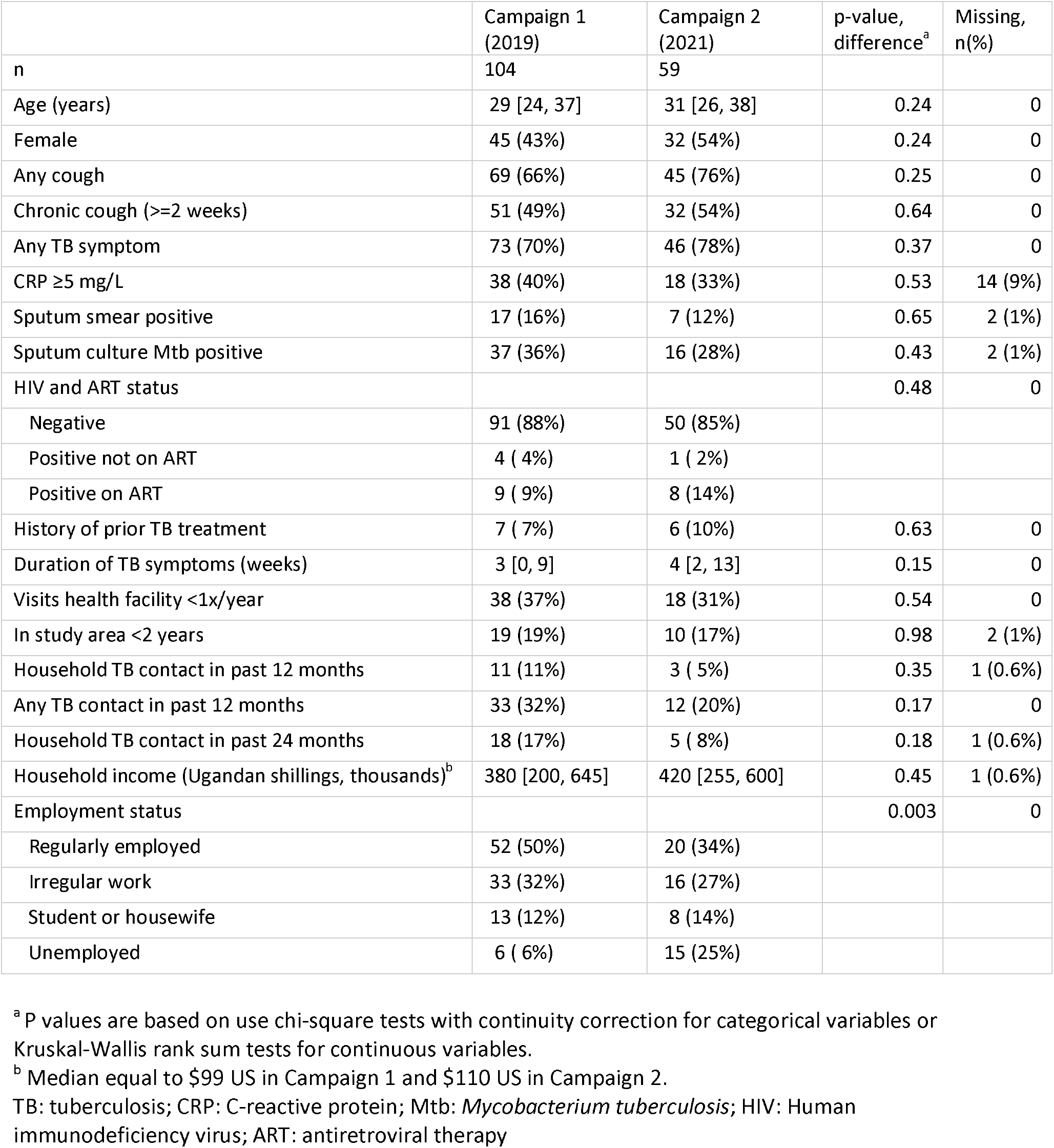
Characteristics of individuals diagnosed with prevalent TB in an urban Ugandan community during two rounds of screening in 2019 (campaign 1) and 2021 (campaign 2)

**Table 3:**
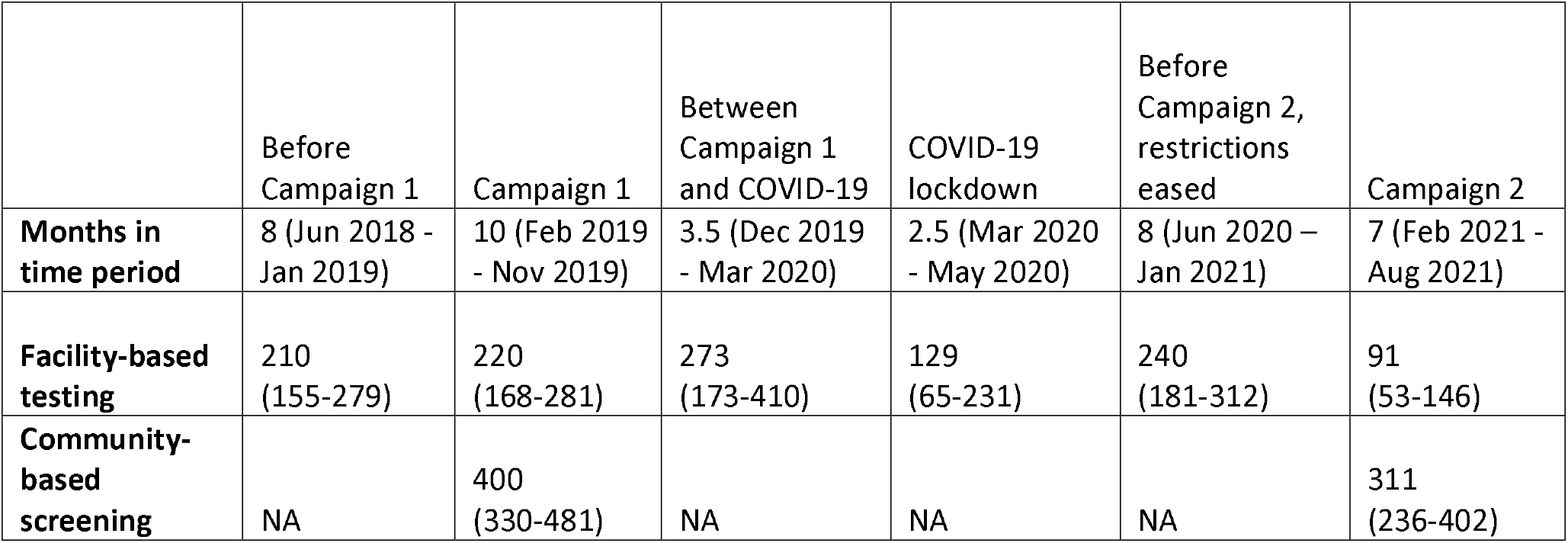
Annualised rates (95% confidence intervals) of facility-based TB diagnosis and community-based tuberculosis detection, per 100,000 estimated adult/adolescent study area residents.

There was no observable change over time in the characteristics of health facility-diagnosed cases (Table S6).

### TB-Related Population Characteristics

Comparing people who tested Xpert-positive in the second campaign to the first, no large or statistically significant differences were observed in disease severity (e.g. prevalence of symptoms, abnormal CRP, smear or culture positivity), symptom duration, prior TB history, HIV co-prevalence, contact with the health care system, or recent migration to the study area (Table 2, Table S5).

In the second campaign, fewer TB-negative community controls reported cough (19% versus 40%) and chronic cough (10% versus 18%) compared to the first campaign (Table S5). Among community cases, however, the prevalence of reported cough increased by a statistically nonsignificant amount (76% versus 66%; 54% versus 49% for chronic cough) (Table 2). Unemployment was likewise higher during the second campaign among cases (25% versus 6%, Table 2), but not community controls (Table S5).

The proportion of people reporting a TB contact within the past year fell over time; reductions were seen among people diagnosed with TB in the community (32% vs 20%, p=0.17, Table 2), community controls (23% vs 1%, p=0.0001, Table S5), and people diagnosed with TB through routine care (39% vs 23%, p=0.04, Table S6).

## Discussion

These two community-wide active-case-finding campaigns in the same high-TB-burden Ugandan community, timed just before and just after the first year of the COVID pandemic, offer insight into the combined effects of the pandemic response and active case-finding efforts on the burden of prevalent TB. The prevalence of Xpert-positive TB among screening participants was 45% lower in the second campaign, despite similar rates of routine notification immediately preceding each campaign. These findings indicate that a substantial reduction in TB burden was achieved in this community over a two-year period, in the context of intensive community-based case finding and any potential impacts of the COVID-19 pandemic response.

The two campaigns used the same methodology, achieved similar population coverage, and found similar characteristics among people diagnosed with TB in both campaigns. As such, it seems likely that the lower TB prevalence observed during the second campaign represents a true reduction in community TB burden, rather than a difference in data collection between the two campaigns (e.g., different populations participating). Our intensive community-based case-finding activities increased case detection (more than doubling the rate of TB diagnosis during the first campaign compared to the baseline rate), removing prevalent TB from the population and preventing new transmission-related cases. However, a 45% reduction in prevalence -- comparable to the 41% reduction in prevalence seen after six rounds of symptom-based case-finding in Zimbabwe [11] and the 44% reduction seen after three rounds of population-wide sputum screening in rural Vietnam [10] – is unlikely to be solely by one case-finding campaign, particularly considering its 35% population coverage and subsequent 35% estimated population between the two campaigns. Rather, the observed prevalence reduction was likely multicausal.

Besides directly detecting TB, our case-finding campaigns also contained a strong community mobilisation component, and we observed sustained rates of care-seeking and TB diagnosis during all but the most restrictive period of pandemic lockdowns. As observed in other case-finding studies [20], increased TB awareness resulting from the study may have encouraged people to seek care for TB symptoms even if not directly screened, or increased clinicians’ index of suspicion for TB. In addition, the regular presence of research staff at local health facilities, and their inquiries about patients who had tested positive for TB and not yet started treatment, may have improved treatment initiation for patients diagnosed with TB through routine care.

These “indirect” effects of our intervention likely contributed to the lower observed TB prevalence during the second campaign.

Along with these effects of our intervention, external factors the COVID-19 pandemic response may also partially explain our findings. The pandemic is generally assumed to have increased transmission of *M. tuberculosis*, through disruptions in care and resulting increases in disease prevalence. However, our study community experienced only a brief reduction in TB case notifications during the pandemic, such that any corresponding increase in transmission might have been small, and social distancing and other effects (e.g., masking, altered contact patterns) could counteract this effect. Finally, TB prevalence in Uganda (which has been measured in a national prevalence survey only once) could be declining more broadly. Antiretroviral therapy and TB preventive treatment have been scaled up substantially for people with HIV, and annual notifications have increased >50% since 2015 [1], coinciding with the rollout of rapid molecular diagnostics (particularly in Kampala) [21] and other efforts to strengthen systems for TB awareness, diagnosis, and treatment [22]. Although official World Health Organization estimates are that TB incidence increased in Uganda between 2019 and 2021 [1], it is possible that these investments in TB and HIV control have reduced TB both nationally and among our study participants.

Our study has important limitations. First, as indicated above, we cannot discern the degree to which observed changes in TB burden reflect effects of our interventions versus secular (including pandemic-related) trends. However, our two in-depth prevalence measurements that flanked the primary pandemic response provide a unique window on the combined impact of intensive active case-finding and the pandemic. Second, even though the populations who participated in our two campaigns were similar in almost all measured characteristics, the COVID-19 pandemic may nonetheless have changed patterns of participation in less apparent ways. In particular, we may have missed reductions in routine TB diagnosis during the COVID-19 period, if mobility restrictions made residents of our study area more likely to seek care at local health facilities (where we were tracking notifications) than at more distant facilities (where we were not). Third, our use of a single sputum Xpert test (rather than a combined microbiological standard based on multiple specimens) likely resulted in both false-negative and false-positive results – and the clinical relevance of “trace” positive results in community settings remains uncertain. However, given that both campaigns used the same specimen collection and testing methods, misclassification of TB status would likely bias our estimated prevalence ratios toward the null. Furthermore, we observed substantially fewer trace-positive results in the second campaign. This finding would be unlikely if such results reflect pure false-positives, and it might instead reflect diagnosis and treatment of people with minimal [23], cyclic [24], or chronic [25] forms of TB that otherwise would have been likely to persist until the second campaign.

In summary, we performed two intensive case-finding campaigns in a well-defined community of Kampala, Uganda, in 2019 and 2021. We observed 45% lower TB prevalence among participants in the second campaign, despite the disruptive effects of the COVID-19 pandemic. This lower prevalence likely reflects the direct effects of the first campaign as well as its indirect effects on TB care-seeking and diagnosis patterns, changes in contact patterns due to COVID-related disruptions, and country-wide secular trends. Our findings offer hope that, even in the context of the COVID-19 pandemic, substantial reductions in TB burden can be achieved through the direct and indirect effects of community-wide case-finding.

## Data Availability

All data produced in the present study are available upon reasonable request to the authors

