## Supplement for "Decline in prevalence of tuberculosis following an intensive case-finding campaign and the COVID-19 pandemic in an urban Ugandan community"

### Supplemental methods

#### Supplemental text 1: Control and contact investigation

During the two active case-finding campaigns, we recruited household and close non-household contacts of both facility-diagnosed and activity-diagnosed TB index cases, for detailed investigation that including sputum Xpert testing for adults/adolescents who had not been screened for TB in the past 4 weeks. These contact-specific sputum tests were not included in our primary estimate of prevalence. Based on contemporary national guidelines, close contacts were only referred for preventive therapy if they were under five years old or living with HIV; we did not verify whether these contacts initiated or completed TPT.

Of 371 contacts investigated during the two campaigns, 183 were adults/adolescent eligible for sputum Xpert testing outside of our routine screening activities; 3 (1.6%) of these tested positive (Figure 1). Another 89 were eligible for preventive therapy referral on the basis of age <5 (68 contacts) or HIV status (21 contacts).

Xpert-negative community controls underwent the same evaluation as cases, including sputum culture. If a control's sputum culture was positive for M. tuberculosis, that individual was reclassified as having TB (though counted as Xpert-negative for our primary prevalence estimate), and additional controls were selected for the original and newly diagnosed cases.

#### Supplemental text 2: Management of trace Xpert Ultra results

One characteristic of Xpert Ultra is the "trace MTB detected" result, indicating a very small quantity of DNA [1]. Of the individuals with trace-positive results in either campaign who participated in further data collection and had interpretable culture results, 12% (10/84) had M. tuberculosis on culture of a subsequent sputum specimen.

During the first campaign, all individuals with trace results were referred to a health center, where most received TB treatment prescribed by non-study providers. During the second campaign, reflecting updated knowledge of the frequency of negative cultures in this group [2], treatment referral for participants with trace-positive results was deferred until additional diagnostic or clinical data confirmed a TB diagnosis. In both campaigns, individuals with trace results were included in the primary estimate of TB prevalence and recruited for more detailed investigations, based on part on the observations that such results are associated with the incidence of TB at a population level [3] and with subtle clinical abnormalities at an individual level [2].

1. Chakravorty S, Simmons AM, Rowneki M, Parmar H, Cao Y, Ryan J, Banada PP, Deshpande S, Shenai S, Gall A, Glass J, Krieswirth B, Schumacher SG, Nabeta P, Tukvadze N, Rodrigues C, Skrahina A, Tagliani E, Cirillo DM, Davidow A, Denkinger CM, Persing D, Kwiatkowski R, Jones M, Alland D. The New Xpert MTB/RIF Ultra: Improving Detection of Mycobacterium tuberculosis and Resistance to Rifampin in an Assay Suitable for Point-of-Care Testing. *mBio* 2017; 8.

2. Kendall EA, Kitonsa PJ, Nalutaaya A, Erisa KC, Mukiibi J, Nakasolya O, Isooba D, Baik Y, Robsky KO, Kato-Maeda M, Cattamanchi A, Katamba A, Dowdy DW. The Spectrum of Tuberculosis Disease in an Urban Ugandan Community and Its Health Facilities. *Clin Infect Dis* 2021; 72: e1035–e1043.

3. Dorman SE, Schumacher SG, Alland D, Nabeta P, Armstrong DT, King B, Hall SL, Chakravorty S, Cirillo DM, Tukvadze N, Bablishvili N, Stevens W, Scott L, Rodrigues C, Kazi MI, Joloba M, Nakiyingi L, Nicol MP, Ghebrekristos Y, Anyango I, Murithi W, Dietze R, Lyrio Peres R, Skrahina A, Auchynka V, Chopra KK, Hanif M, Liu X, Yuan X, Boehme CC, et al. Xpert MTB/RIF Ultra for detection of Mycobacterium tuberculosis and rifampicin resistance: a prospective multicentre diagnostic accuracy study. *Lancet Infect Dis* 2018; 18: 76–84.

#### Supplemental text 3: Estimation of population and people evaluated for presumptive TB

Estimates of the community population size were based on study staff's enumeration of study area residences and residents during each campaign. For residences on which occupancy information was unavailable, we assumed similar rates of vacancy and household size as for residences on which this information was documented.

To estimate the number of study area residents who underwent routine TB diagnostic evaluation during the study, we tallied the patients in the four local facilities' presumptive TB registers, by documented residence (within/outside the study area) and the date range of each register page. One facility's presumptive TB register was missing for a one-year period in 2018-2019; for estimating total TB evaluations, we estimated that the monthly number of patients who sought care during this period was the same as the monthly average during periods with available data.

#### Supplemental text 4: COVID-related restrictions in study area

Uganda imposed restrictions on international travel in early March 2020. Two lockdowns in March-May 2020 and in June-August 2021 (lasting 63 and 42 days respectively, then gradually eased) restricted most types of public and private gatherings as well as nonessential public and private transport. Other restrictions were more prolonged. Most schools were closed for nearly two years from March 2020 to January 2022. A 7pm evening curfew was in place from March 2020 to January 2022 with restrictions on transportation and business operations. Large gatherings (e.g. weddings, church services) were prohibited from March 2020 to August 2021.

#### Supplemental text 5: Obtaining sputum samples for Xpert testing

As previously described for the first campaign, we coached all consenting participants on how to provide the best quality sputum that they were able. We ultimately obtained a sputum sample of at least 1 ml from 99.5% of consenting participants (23879/24010). Laboratory personnel rated 75% of samples as salivary and 54% had a volume of only 1ml, but the proportion of Xpert results that were positive was only modestly lower among salivary versus other sputum qualities (0.5% vs 1.1%) and marginally lower among 1ml versus higher-volume specimens (0.6% vs 0.7%).

#### Table S1. Primary and secondary prevalence estimation methods

| | Numerator | Denominator | Confidence interval | Notes |
|---|---|---|---|---|
| **Primary estimate: Community prevalence among screened adults** | Individuals with a positive (including trace) Xpert result in community-wide TB screening, Feb-Nov 2019.* | Individuals with a valid (positive, negative, or trace) Xpert result in community-wide TB screening, Feb-Nov 2019 | Exact binomial 95%CI (R package 'binom') | Excludes tests repeated during contact investigation, or tested during contact investigations and control enrollments that occurred in December 2019 (i.e., that lagged behind the end of the community-wide testing period) |
| **Age- and sex-adjusted adult prevalence** | Proportion of Xpert tests positive as for primary estimate, but stratified by age (15-25, 25-35, 35-45, 45-55, or >55 years) and adjusted for population proportion among residents enumerated during door-to-door case-finding visits | Valid Xpert tests as for primary estimate, adjusted for population proportion in age strata 15-25, 25-35, 35-45, 45-55, and >55 years among residents enumerated during door-to-door case-finding visits | Byar's method [a] with Dobson method adjustment [b] | Calculated using R package PHEindicatormethods |
| **Adult prevalence excluding culture-negative trace-positives** | As for primary estimate, but subtracting estimated number of trace-positive sputum that would be culture negative | As for primary estimate, but subtracting estimated number of trace-positive, culture-negative sputa (excluding these individuals from analysis) | Exact binomial 95%CI | The proportion culture-negative was determined among trace positive sputa with a valid sputum culture result, then applied to all trace positive sputa. |
| **Total proportion of adult population diagnosed with TB during case-finding period** | All adults diagnosed with pulmonary TB through community-based case finding (including contacts and controls) at local health facilities, from Feb through Nov 2019 | Total estimated adult population of study area | Exact binomial 95%CI | Households lacking a count of adult residents were assumed to have the same average size as households for which a household member or neighbor provided a count. |

[a] Breslow NE, Day NE.Statistical methods in cancer research, volume II: The design and analysis of cohort studies. Lyon: International Agency for Research on Cancer, World Health Organisation; 1987. [b] Dobson A et al. Confidence intervals for weighted sums of Poisson parameters. Stat Med 1991;10:457-62.

* To maintain consistent sampling frames for TB-positive and TB-negative individuals, prevalence estimates were based on residence as reported at time of screening (even if incorrectly reported), but case or control eligibility required a confirmed residence within the study area.

### Supplemental Results

#### Figure S1: Detailed flowchart of first community campaign

(adapted/expanded from Kendall et al., CID 2020)

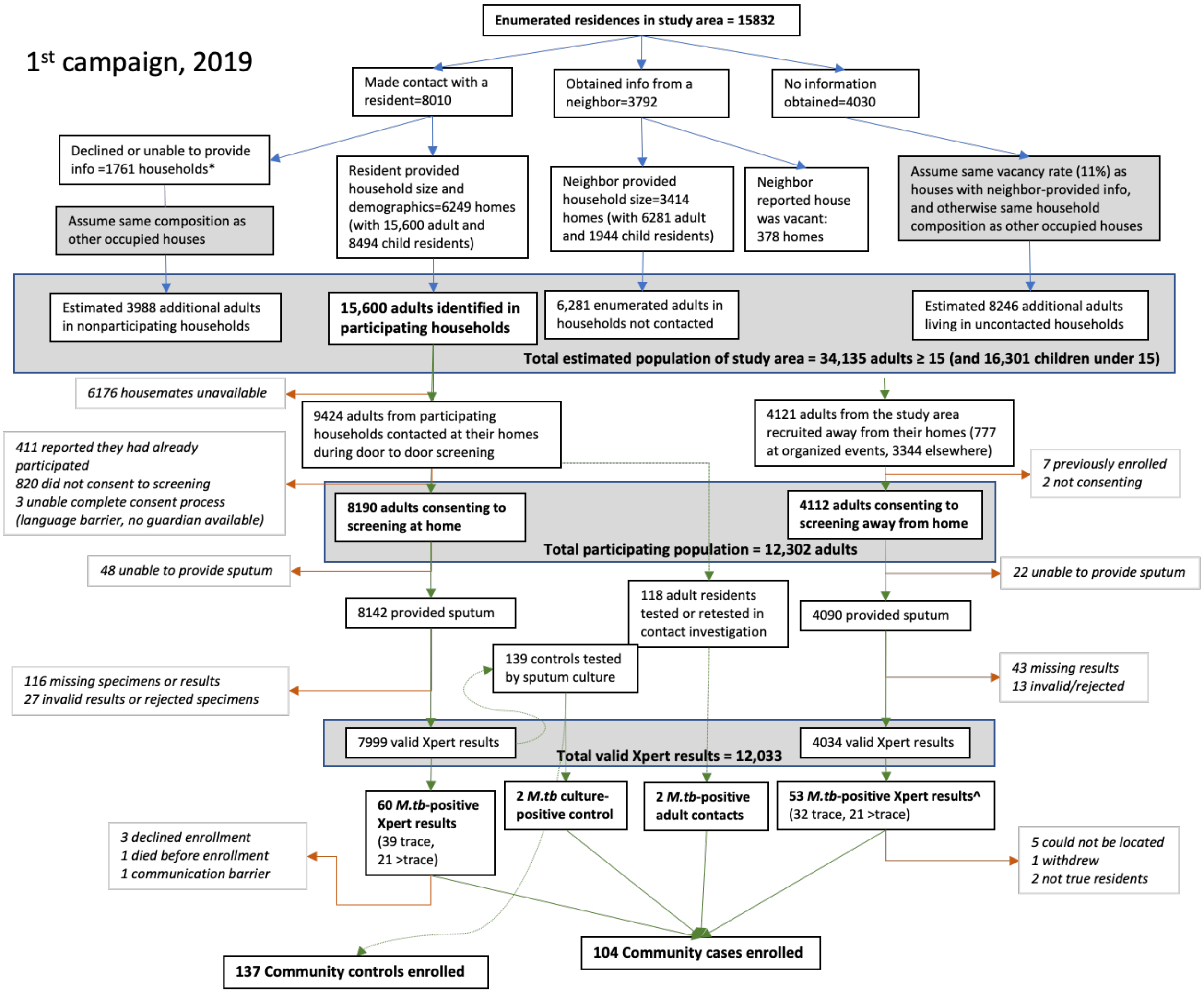

### Figure S2: Detailed flowcharts of second community campaign

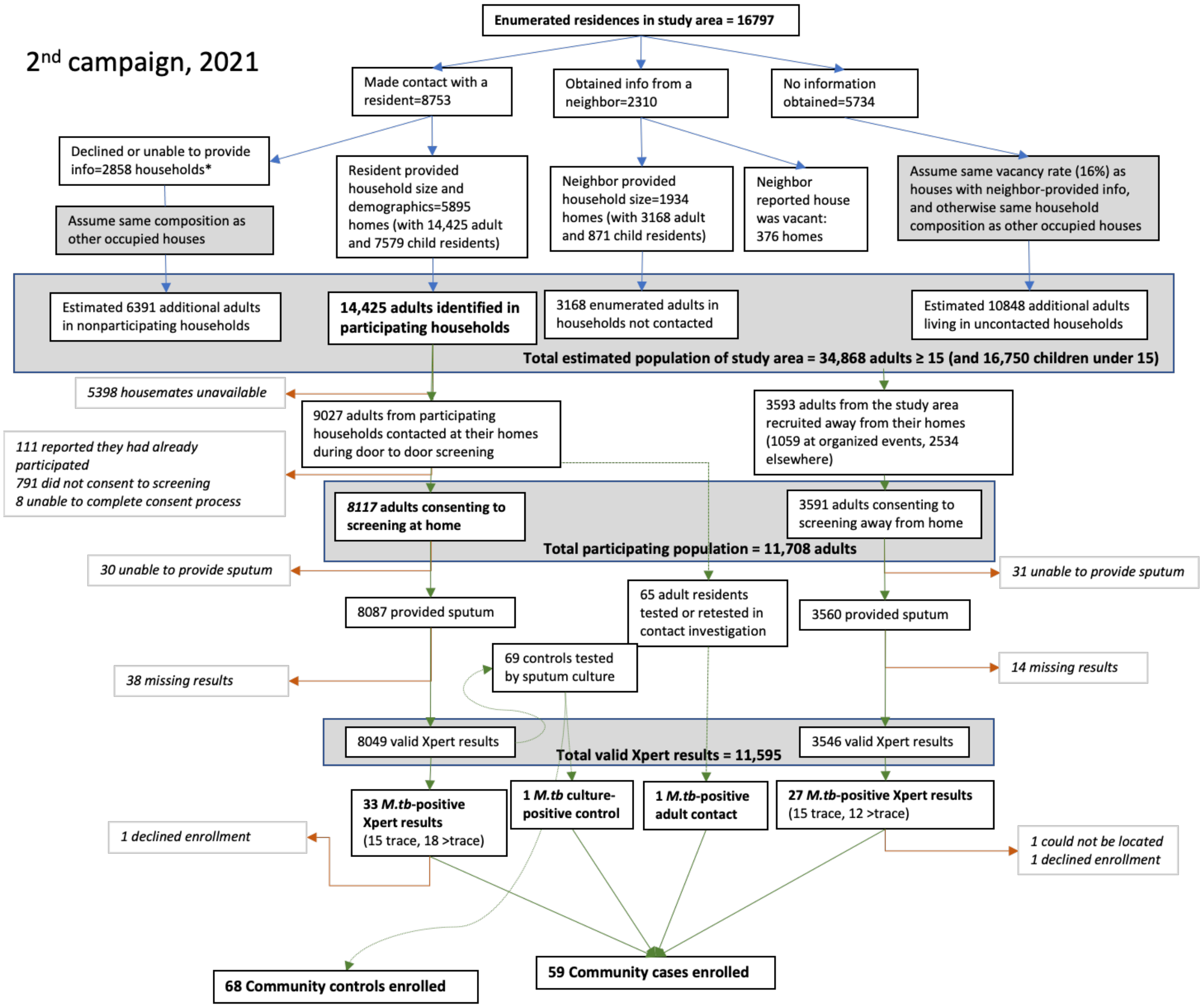

#### Figure S3: Comparison of household occupancy and participation

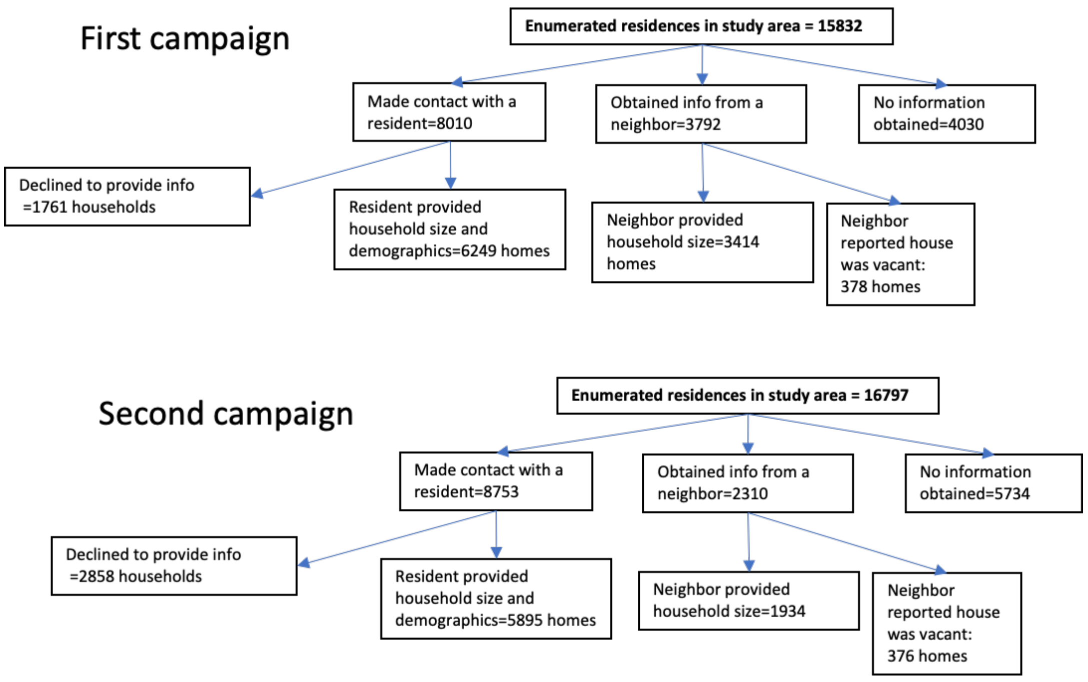

Figure S4: Age and sex composition of the enumerated population (top) and screened participants (bottom) **were consistent across two community-based tuberculosis case-finding campaigns. The age/sex composition of participants resembled that of the overall population except for slight overrepresentation of younger women and underrepresentation of older men.**

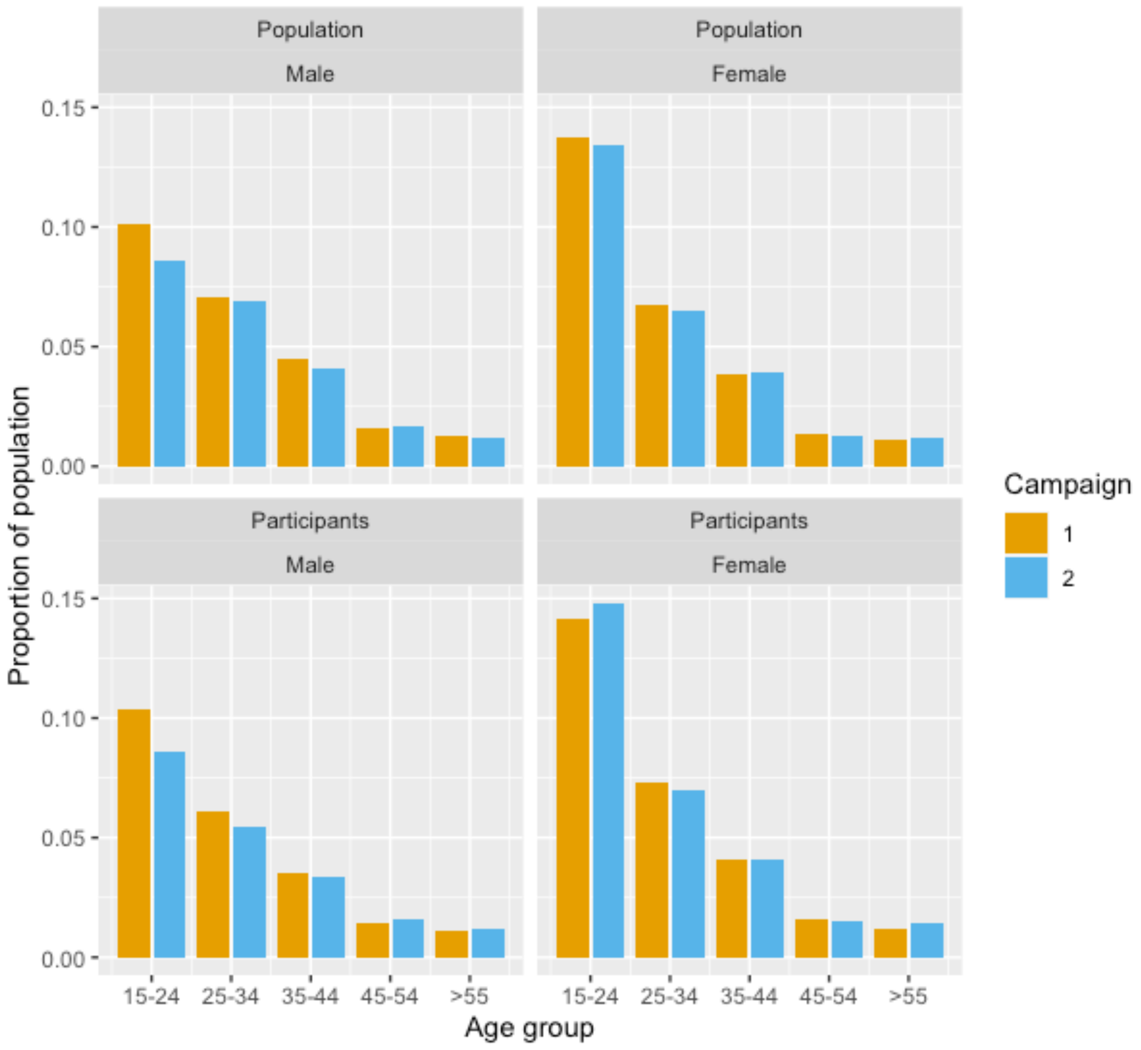

Figure S5: Prevalence and participation by zone and screening location. The left panel shows the prevalence of sputum Xpert-positive TB among participants within each zone (with 95% binomial confidence intervals), and the right panels show total screening participants from each zone, stratified by screening location (home vs other). Zones are arranged in descending order according to TB prevalence among participants from that zone (pooled across the two campaigns). This depiction allows verification that higher overall prevalence in campaign 1 was not driven by higher overall or away-from-home participation in the zones with either the highest prevalence or the largest number of people screened. Zones that contributed <220 participants across both campaigns (<1% of the total sample) are omitted from the figure.

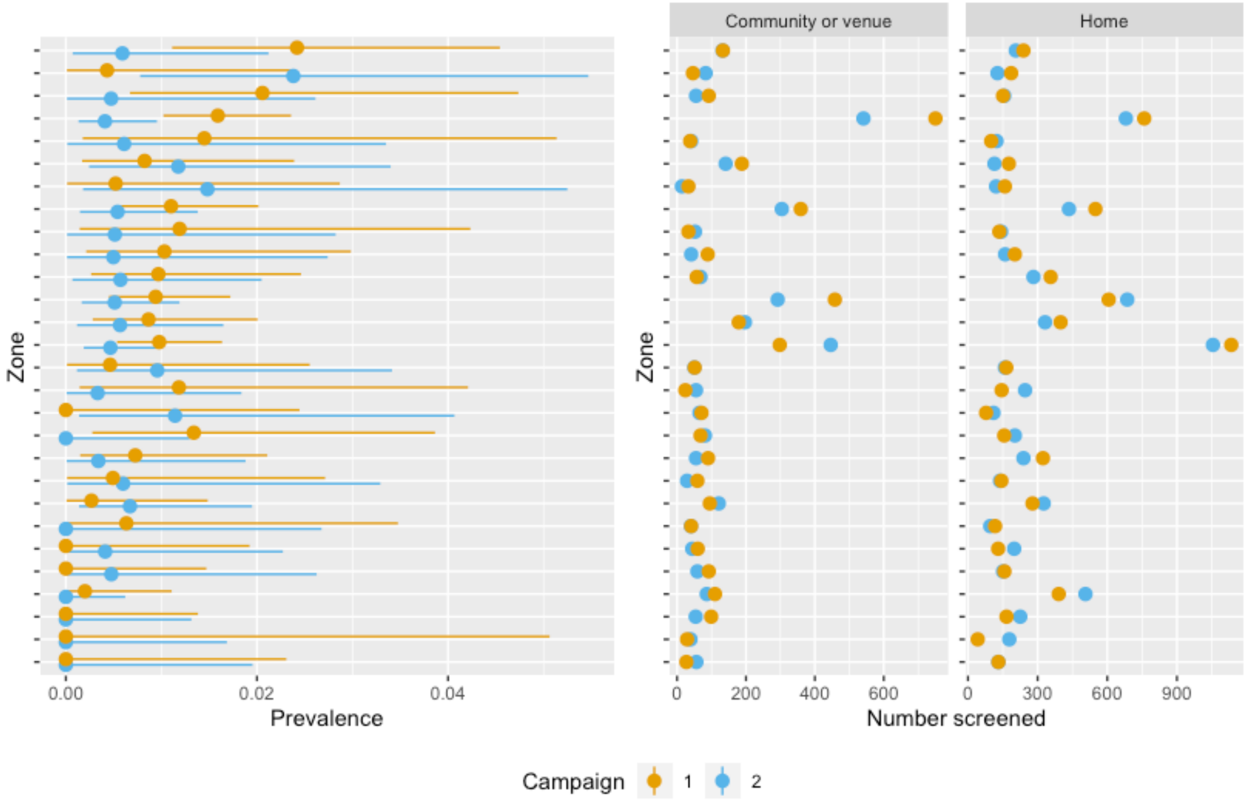

#### Supplemental text 6: Estimating repeated participation

Repeated Participation Methods

Estimates of repeat participation were generated from iris ID data by considering the number of screening participants in each campaign ($S_1$, $S_2$), the number of screening participants with iris IDs recorded in each campaign ($I_1$, $I_2$), the number of matching iris IDs that were recorded in both campaigns ($I_{12}$), and the estimated size of the eligible population in each campaign ($P_1$ and $P_2$). Here, $S_{12}$ refers to the number of people who were screened in both campaigns, and $P_{12}$ refers to the number of people who were in the eligible population during both campaigns.

We made two assumptions:

1. Each campaign's screening participants were a random sample of study area residents at that time: $S_{12} = P_{12} \cdot S_1/P_1 \cdot S_2/P_2$
2. Missing iris IDs were missing at random: $I_{12} = S_{12} \cdot I_1/S_1 \cdot I_2/S_2$

Under these assumptions, we estimated the proportion of campaign-2 screening participants that had also participated in campaign 1 as $S_{12}/S_2 = I_{12}S_1 / I_1I_2$ , and the number who had participated in both campaigns as $I_{12}S_1S_2 / I_1I_2$.

Similarly, we estimated the proportion of the campaign-2 eligible population who had been eligible residents during campaign 1 as $P_{12}/P_2 = S_{12}P_1 / S_1S_2 = I_{12}P_1 / I_1I_2$, and the number eligible for both phases as $I_{12}P_1 P_2 / I_1I_2$. Of the remaining population estimated to have been eligible during campaign 2 but not campaign 1, we estimated new 15-year-olds based on the proportion of the enumerated population who were 15 or 16 years old during campaign 2, and assumed that the rest of the newly eligible population had immigrated within the past two years.

Finally, we estimated what proportion of community cases found during campaign 2 had also participated in campaign 1, as $n(C_2 \cap S_1)/ C_2 = n(C_2 \cap I_1)\ (S_1/I_1)\ /C_2$ , and similarly, the proportion of campaign 1 cases who also participated in campaign 2 as $n(C_1 \cap I_2)\ (S_2/I_2)\ /C_1$.

Repeated Participation Results

Iris IDs were obtained from 71% of participants in the first campaign and 44% of participants in the second campaign (when many participants raised concerns related to social distancing). During the second campaign, 791 of 5183 IDs (15%) matched an ID from the first campaign. We estimated that 2500 individuals (comprising 20% of campaign 1 participants and 22% of campaign 2 participants) participated in both campaigns (Table S2). After accounting for missing IDs, aging, and migration, we estimated for each campaign that 33-37% of the participants eligible for the other campaign had participated in the other campaign as well (Table S2). A similar level of repeated participation was estimated for the people diagnosed with TB in either campaign, although sample sizes were small.

Estimates of repeat participant were consistent when restricted to participants with positive campaign 2 screening results (2 of 20 such IDs were matched to campaign-1 screening participant, and after adjusting for missing iris IDs we estimated that 14% had participated in campaign 1); participants with positive campaign 1 screening results (8 of 80 matched; estimated 23% repeat participation in campaign 2); participants who were enrolled as campaign 2 community cases (10 of 47 matched; adjusted estimate that 30% had participated in campaign 1); participants who were enrolled in campaign 2 as TB-negative community controls (9 of 54 matched; adjusted estimate that 23% had participated in campaign 1); and health facility cases diagnosed after the campaign 1 (6 of 59 matched; estimated 14% had participated in campaign 1, with two of seven identifiable matches having had positive screening results and received treatment during campaign 1). Self-reported rates of campaign 1 screening participation were also consistent with iris-based estimates: 26% (15/59) of campaign 2 community cases and 25% (18/68) of campaign 2 community controls.

Using a similar iris-ID-based approach and adjusting for missing IDs, we estimated that 59% of the eligible campaign-2 population had been eligible during campaign 1 (Table S1). Another 6% of the eligible population were fifteen and sixteen-year-olds who were newly eligible based on age. This suggests that 35% of the campaign-2 population were new migrants within the past 2 years. Enrolled community case and control participants may have had stronger ties to the study area than the average resident, but they also reported high rates of migration, with 18% (65/366) reporting that they were new to the study area within the past 1.5 years; this did not differ between campaigns.

#### Table S2: Iris-based estimates of overlap and turnover in screened population

| | **Campaign 1** | **Campaign 2** |
|---|---|---|
| **Estimated total eligible population of study area** | 34135 | 34869 |
| **Participants consenting to screening (% of population)** | 12302 (36%) | 11708 (34%) |
| **Raw iris identification data (% of screened)** | | |
| *Without iris ID* | *3510 (29%)* | *6525 (56%)* |
| *With iris, not matched across campaigns* | *8001 (65%)* | *4392 (38%)* |
| *With iris, matched across campaigns* | *791 (6%)* | *791 (7%)* |
| **Estimates of participation across campaigns *(% of screened)*:** | | |
| *Participated in both campaigns* | *2500 (20%)* | *2500 (22%)* |
| *Eligible for both, but did not participate in campaign 1* | - | *4437 (38%)* |
| *New 15-year-olds between campaigns 1 and 2* | - | *717 (6%)* |
| *New immigrants between campaigns 1 and 2* | - | *4054 (35%)* |
| *Eligible for both, but did not participate in campaign 2* | *4946 (40%)* | - |
| *Left area or died before phase 2* | *4856 (39%)* | - |

#### Table S3: Estimated TB prevalence ratios, Campaign 2 (2021) versus Campaign 1 (2019), after multivariable adjustment

| | **Campaign (2 vs 1)** | **Age** | **Female** | **Parish of residence** | **Home screening location** |
|---|---|---|---|---|---|
| Unadjusted | 0.55 (0.40 - 0.75) | NA | NA | NA | NA |
| Adjusted for the indicated variables | 0.56 (0.41 - 0.76) | 1 (0.99 - 1.01) | 0.64 (0.48 - 0.87) | NA | NA |
| | 0.56 (0.41 - 0.76) | 1 (0.99 - 1.01) | 0.64 (0.48 - 0.87) | 1 (0.84 - 1.19) | NA |
| | 0.56 (0.41 - 0.77) | 1.01 (1 - 1.02) | 0.74 (0.54 - 1.01) | 1.18 (0.99 - 1.4) | 0.63 (0.46 - 0.86) |
| | 0.76 (0.47 - 1.21) | 1.02 (1 - 1.03) | 0.48 (0.29 - 0.79) | 1.09 (0.84 - 1.43) | 0.7 (0.44 - 1.13) |

#### Figure S6: Prevalence decreased in nearly all age and sex strata, but the largest reductions were seen among men.

Prevalence estimates are shown with 95% binomial confidence intervals.

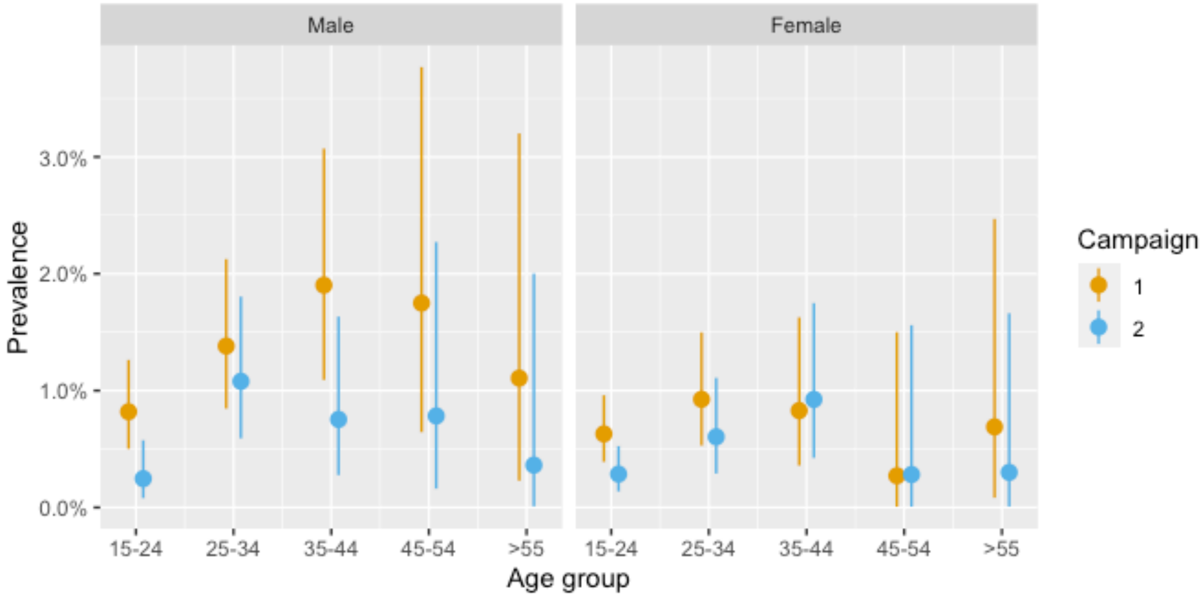

Figure S7: Despite a higher proportion of trace results in campaign 1 versus campaign 2, there was no consistent pattern of lower semiquantitative Xpert Ultra results in campaign 1.

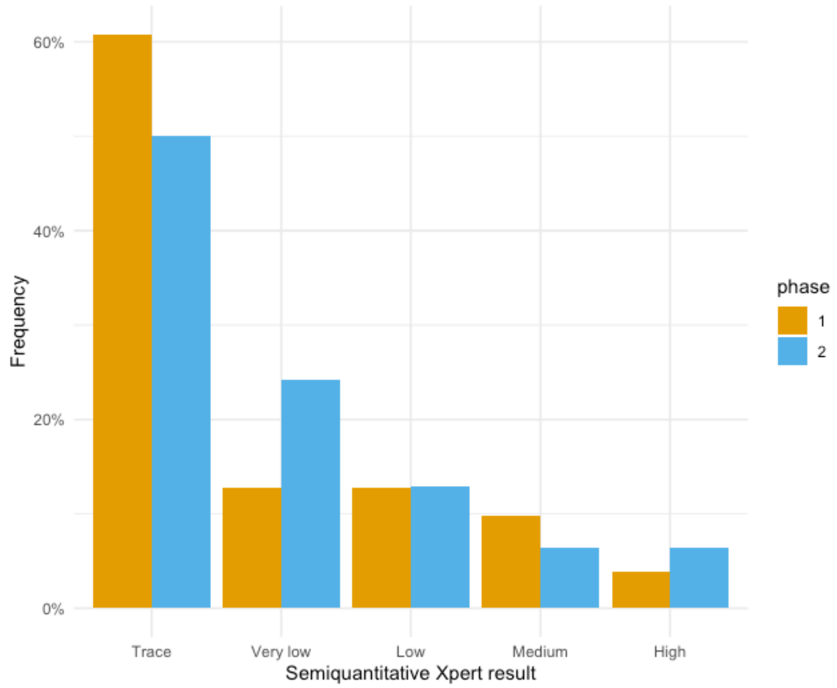

Table S4: Characteristics of people with positive TB screening results in the first (2019) versus second (2021) community screening campaigns, limited to people whose positive Xpert results were greater than trace or confirmed by culture.

| | Campaign 1 | Campaign 2 | p | % Missing |
|---|---|---|---|---|
| **n** | 49 | 35 | | |
| **Age (years)** | 31 [24, 41] | 32 [26, 38] | 0.913 | 0 |
| **Female** | 14 (29%) | 14 (40%) | 0.389 | 0 |
| **Any cough** | 42 (86%) | 28 (80%) | 0.692 | 0 |
| **Chronic cough (>=2 weeks)** | 35 (71%) | 23 (66%) | 0.75 | 0 |
| **Any TB symptom** | 40 (82%) | 31 (89%) | 0.575 | 0 |
| **CRP ≥5 mg/L** | 21 (50%) | 13 (41%) | 0.571 | 11.9 |
| **Sputum smear positive** | 17 (35%) | 7 (21%) | 0.251 | 1.2 |
| **Sputum culture Mtb positive** | 37 (76%) | 16 (47%) | 0.015 | 1.2 |
| **HIV and ART status** | | | 0.379 | 0 |
| **Negative** | 43 (88%) | 29 (83%) | | |
| **Positive not on ART** | 3 ( 6%) | 1 ( 3%) | | |
| **Positive on ART** | 3 ( 6%) | 5 (14%) | | |
| **History of prior TB treatment** | 3 ( 6%) | 4 (11%) | 0.64 | 0 |
| **Duration of TB symptoms (weeks)** | 4 [2, 16] | 4 [2, 16] | 0.837 | 0 |
| **Visits health facility <1x/year** | 19 (39%) | 9 (26%) | 0.309 | 0 |
| **In study area <2 years** | 7 (14%) | 8 (23%) | 0.47 | 0 |
| **Household TB contact in past 12 months** | 4 ( 8%) | 3 ( 9%) | 1 | 1.2 |
| **Any TB contact in past 12 months** | 18 (37%) | 9 (26%) | 0.407 | 0 |
| **Household TB contact in past 24 months** | 7 (15%) | 4 (11%) | 0.928 | 1.2 |
| **Household income (Ugandan shillings, thousands)** | 300 [167, 500] | 320 [200, 550] | 0.398 | 0 |
| **Employment status** | | | 0.017 | 0 |
| **Regularly employed** | 24 (49%) | 11 (31%) | | |
| **Irregular work** | 20 (41%) | 12 (34%) | | |
| **Student or homemaker** | 4 ( 8%) | 4 (11%) | | |
| **Unemployed** | 1 ( 2%) | 8 (23%) | | |

Table S5: Characteristics of TB-negative community controls enrolled during the first (2019) versus second (2021) community screening campaigns

| | Campaign 1 | Campaign 2 | | % Missing |
|---|---|---|---|---|
| n | 137 | 68 | | |
| Age (years) | 26 [22, 35] | 26 [21, 33] | 0.466 | 0 |
| Female | 92 ( 67%) | 49 ( 72%) | 0.58 | 0 |
| Any cough | 55 ( 40%) | 13 ( 19%) | 0.004 | 0 |
| Chronic cough (>=2 weeks) | 24 ( 18%) | 7 ( 10%) | 0.249 | 0 |
| Any TB symptom | 52 ( 38%) | 22 ( 32%) | 0.527 | 0 |
| CRP ≥5 mg/L | 22 ( 19%) | 4 ( 8%) | 0.12 | 17.6 |
| Sputum smear positive | 1 ( 1%) | 3 ( 4%) | 0.221 | 3.4 |
| Sputum culture Mtb positive | 0 ( 0%) | 0 ( 0%) | NA | 3.4 |
| HIV and ART status | | | 0.339 | 0 |
| Negative | 123 ( 90%) | 65 ( 96%) | | |
| Positive not on ART | 1 ( 1%) | 0 ( 0%) | | |
| Positive on ART | 13 ( 9%) | 3 ( 4%) | | |
| History of prior TB treatment | 4 ( 3%) | 0 ( 0%) | 0.375 | 0 |
| Duration of TB symptoms (weeks) | 0 [0, 4] | 0 [0, 2] | 0.103 | 0 |
| Visits health facility <1x/year | 31 ( 23%) | 21 ( 31%) | 0.268 | 0 |
| In study area <2 years | 24 ( 18%) | 12 ( 18%) | 1 | 0 |
| Household TB contact in past 12 months | 5 ( 4%) | 0 ( 0%) | 0.265 | 0 |
| Any TB contact in past 12 months | 32 ( 23%) | 1 ( 1%) | <0.001 | 0 |
| Household TB contact in past 24 months | 11 ( 8%) | 0 ( 0%) | 0.038 | 0 |
| Household income (Ugandan shillings, thousands) | 400 [200, 500] | 425 [250, 725] | 0.068 | 0 |
| Employment status | | | 0.726 | 0 |
| Regularly employed | 72 ( 53%) | 35 ( 51%) | | |
| Irregular work | 18 ( 13%) | 11 ( 16%) | | |
| Student or homemaker | 39 ( 28%) | 16 ( 24%) | | |
| Unemployed | 8 ( 6%) | 6 ( 9%) | | |

Table S6: Characteristics of health facility cases diagnosed before versus after TB screening occurred in their neighborhoods in 2019

| | Before | After | p | % Missing |
|---|---|---|---|---|
| **n** | 65 | 97 | | |
| **Age (years)** | 32 [28, 40] | 35 [27, 43] | 0.259 | 0 |
| **Female** | 20 ( 31%) | 33 (34%) | 0.794 | 0 |
| **Any cough** | 64 ( 98%) | 89 (92%) | 0.14 | 0 |
| **Chronic cough (>=2 weeks)** | 62 ( 95%) | 82 (85%) | 0.058 | 0 |
| **Any TB symptom** | 65 (100%) | 93 (96%) | 0.254 | 0 |
| **CRP ≥5 mg/L** | 52 ( 81%) | 69 (78%) | 0.822 | 6.1 |
| **Sputum smear positive** | 35 ( 56%) | 38 (41%) | 0.101 | 3.7 |
| **Sputum culture Mtb positive** | 46 ( 73%) | 56 (60%) | 0.14 | 3.7 |
| **HIV and ART status** | | | 0.13 | 0 |
| **Negative** | 38 ( 58%) | 67 (69%) | | |
| **Positive not on ART** | 6 ( 9%) | 12 (12%) | | |
| **Positive on ART** | 21 ( 32%) | 18 (19%) | | |
| **History of prior TB treatment** | 18 ( 28%) | 17 (18%) | 0.178 | 0 |
| **Duration of TB symptoms (weeks)** | 8 [4, 16] | 8 [4, 16] | 0.8 | 0 |
| **Visits health facility <1x/year** | 19 ( 29%) | 13 (13%) | 0.023 | 0 |
| **In study area <2 years** | 7 ( 11%) | 17 (18%) | 0.337 | 0 |
| **Household TB contact in past 12 months** | 6 ( 9%) | 8 ( 8%) | 1 | 0.6 |
| **Any TB contact in past 12 months** | 25 ( 39%) | 22 (23%) | 0.039 | 0.6 |
| **Household TB contact in past 24 months** | 11 ( 17%) | 13 (13%) | 0.664 | 0.6 |
| **Household income (Ugandan shillings, thousands)** | 380 [200, 620] | 260 [150, 400] | 0.043 | 0 |
| **Employment status** | | | 0.002 | 0 |
| **Regularly employed** | 36 ( 55%) | 28 (29%) | | |
| **Irregular work** | 14 ( 22%) | 47 (48%) | | |
| **Student or homemaker** | 3 ( 5%) | 6 ( 6%) | | |
| **Unemployed** | 12 ( 18%) | 16 (16%) | | |

Table S7: Spectrum of disease observed in health facility and community cases over a 39-month period.

| | Health facility cases | Community cases, Xpert >trace | Community cases, Xpert trace | TB-negative community controls | % Missing |
|---|---|---|---|---|---|
| n | 164 | 66 | 86 | 205 | |
| Age (years) | 35 [27, 41] | 31 [25, 38] | 29 [24, 38] | 26 [22, 35] | 0 |
| Female | 54 (33%) | 22 (33%) | 49 (57%) | 141 (69%) | 0 |
| Any cough | 155 (95%) | 57 (86%) | 52 (60%) | 68 (33%) | 0 |
| Chronic cough (>=2 weeks) | 146 (89%) | 46 (70%) | 35 (41%) | 31 (15%) | 0 |
| Any TB symptom | 160 (98%) | 57 (86%) | 54 (63%) | 74 (36%) | 0 |
| CRP ≥5 mg/L | 123 (80%) | 27 (48%) | 26 (32%) | 26 (15%) | 11.3 |
| Sputum smear positive | 74 (47%) | 23 (35%) | 0 ( 0%) | 4 ( 2%) | 2.8 |
| Sputum culture Mtb positive | 103 (65%) | 37 (57%) | 10 (12%) | 0 ( 0%) | 2.8 |
| HIV and ART status | | | | | 0 |
| Negative | 105 (64%) | 55 (83%) | 76 (88%) | 188 (92%) | |
| Positive not on ART | 18 (11%) | 4 ( 6%) | 1 ( 1%) | 1 ( 0%) | |
| Positive on ART | 41 (25%) | 7 (11%) | 9 (10%) | 16 ( 8%) | |
| History of prior TB treatment | 36 (22%) | 6 ( 9%) | 7 ( 8%) | 4 ( 2%) | 0 |
| Duration of TB symptoms (weeks) | 8 [4, 16] | 4 [2, 16] | 2 [0, 7] | 0 [0, 2] | 0 |
| Visits health facility <1x/year | 32 (20%) | 25 (38%) | 28 (33%) | 52 (25%) | 0 |
| In study area <2 years | 24 (15%) | 10 (15%) | 15 (18%) | 36 (18%) | 0.4 |
| Household TB contact in past 12 months | 14 ( 9%) | 4 ( 6%) | 10 (12%) | 5 ( 2%) | 0.4 |
| Any TB contact in past 12 months | 47 (29%) | 21 (32%) | 21 (24%) | 33 (16%) | 0.2 |
| Household TB contact in past 24 months | 24 (15%) | 7 (11%) | 16 (19%) | 11 ( 5%) | 0.4 |
